# Genetic Associations with Polycystic Ovary Syndrome: The Role of The Mitochondrial Genome; A Systematic Review and Meta-analysis

**DOI:** 10.1101/2022.01.22.22269680

**Authors:** Almira Moosa, Meeladah Ghani, Helen O’Neill

## Abstract

1.

**Background:** Polycystic Ovary Syndrome (PCOS) remains the most common female reproductive endocrine disorder. Genetic studies have predominantly focused on the role of the nuclear genome, whilst the contribution of mitochondrial genetics in PCOS remains largely unknown.

**Aim:** This study aims to systematically evaluate the literature regarding the associations between the mitochondrial genome and PCOS.

**Methods:** A literature search focused on PCOS and mitochondrial genetics was conducted on (1) MEDLINE (2) EMBASE and (3) The Cochrane Library (CENTRAL and Cochrane Reviews). Search results were screened for eligibility, and data involving genetic variants of mitochondrial DNA (mtDNA) was extracted. Quantitative data was presented in forest plots, and where this was not possible, data was analysed in a qualitative manner. Quality of studies was assessed using the Q-Genie tool.

**Results:** Of the 13,812 identified studies, 15 studies were eligible for inclusion, with 8 studies suitable for meta-analysis. Women with PCOS showed higher frequencies of a 9-bp deletion, and aberrant SNPs in the ND5, A6, and 7 tRNA-encoding genes. They also showed lower frequencies of two SNPs in the D-Loop of the genome. Women with PCOS also exhibited significantly lowered mtDNA copy number.

**Conclusion:** Women with PCOS harbour genetic variants in coding and non-coding regions of the mitochondrial genome. This may disrupt the electron transport chain and lead to oxidative stress, causing apoptosis of cells and further genetic damage. However, further studies of higher quality are required to confirm these associations.

## 2. INTRODUCTION

Polycystic Ovary Syndrome (PCOS) remains the most common female reproductive endocrine disorder, affecting 15-20% of women of reproductive age.[1,2] Typically showing signs of anovulation, hyperandrogenemia, dyslipidemia, and insulin resistance (IR), women with PCOS often experience infertility, and are predisposed to a multitude of conditions, including type 2 diabetes mellitus, cardiovascular disease, metabolic syndrome (MetS), and, in later years, endometrial cancer.[2-4] As such, PCOS, although rooted in ovarian pathology, evolves gradually to encompass complex multi-organ and multi-system dysfunction.

The pathogenesis of PCOS remains largely unknown, however two of the most influential factors thought to contribute to the development of PCOS are genetics and oxidative stress. As such, the next step towards understanding this multifaceted disorder is to consider the interface between these two systems, by evaluating the role of the mitochondrial genome in PCOS.

Hyperglycaemic states in women with PCOS, largely attributed to insulin resistance (IR), place the mitochondria under a non-physiological amount of strain, exposing it to high levels of reactive oxygen species (ROS). Its close proximity to the electron transport chain (ETC) therefore leaves mtDNA vulnerable to damage by oxidative stress, particularly given that mtDNA lacks protection from histones and chromatin.[5] Not only is the mitochondrial genome vulnerable to acquiring genetic changes, but it also possesses a lowered capacity to manage them, by way of low-efficiency DNA repair mechanisms,[6] given its dependence on a single DNA polymerase, DNA polymerase-gamma, for both replication and repair.[7,8] Consequently, oxidative stress in PCOS can lead to mtDNA mutagenesis, and an array of metabolic changes. Given the vital role of the organelle, many regions of the mitochondrial genome remain evolutionarily conserved,[9] providing scope for small-scale genetic variants to have large implications in the pathogenesis of this disease.

Whilst it has become accepted that genetic associations exist with PCOS, the majority of research thus far has focused on the nuclear genome, resulting in a sizeable number of studies exploring nuclear-encoded factors and their influence on PCOS. Despite this large pool of literature, however, few definitive associations have been documented, leaving many questions unanswered about the true genetic basis of the disease. Owing to the lack of answers therefore, attention in recent years has been redirected towards the mitochondrial genome, which, up until now, remained largely overlooked.

Following this redirection in research, there has been an increase in the number of studies considering the role of mitochondrial genetics in PCOS, providing new insights into PCOS as a disease, and warranting a systematic review of the evidence on this emerging concept.

As such, this study aims to systematically evaluate the literature regarding the associations between the mitochondrial genome and PCOS. Outcomes of interest for this study are:

- Genetic variants in coding regions of the mitochondrial genome
- Genetic variants in the non-coding region of the mitochondrial genome
- Changes in mitochondrial genetic content
- Quality of included studies

To the authors’ knowledge, this is the first systematic review to consider the mitochondrial genome in the context of PCOS.

## 3. METHODS

This study was conducted in accordance with the PRISMA guidelines,[10,11] and is registered in the International Prospective Register of Systematic Reviews (PROSPERO), with registration number CRD42021267991.

### 3.1 Search Strategy

A literature search was carried out via (1) MEDLINE, (2) EMBASE and (3) The Cochrane Library (CENTRAL and Cochrane Reviews) electronic databases, using subject headings and keywords relating to “polycystic ovary syndrome” and “mitochondrial genetics” (***Supplement 1***). Limits were applied to return only English language and human studies.

Searches were conducted by two reviewers independently, with disagreements being resolved by discussion. Publications spanning all years were considered, with the last search being conducted on 19/07/2021. Citations were exported to EndNote X9 for management, and duplicates were removed. Search results were then screened for eligibility.

Reference lists of included studies and review articles were manually searched to identify articles not sought through electronic searches, and authors were contacted regarding papers with restricted access.

### 3.2 Eligibility Criteria and Study Selection

Two reviewers independently assessed eligibility of studies for inclusion, with discrepancies being resolved by discussion. Publications were initially screened by title and abstract, with seemingly relevant papers then being considered in full.

Studies were included if they met the following criteria:

1. **Population** considered women with a clinical diagnosis of PCOS
2. **Interventions** included genetic investigations of the mitochondrial genome
3. **Comparison** was made to a suitable control group of women without PCOS
4. **Outcomes** included any measures relating to genetic content
5. **Study design** of any type

Thus, studies focused on: animals, the nuclear genome, other reproductive conditions apart from PCOS, and those which did not carry out genetic techniques, were excluded. Review articles, abstract-only articles, reply letters, and inaccessible papers were also excluded.

### 3.3 Data Extraction

Data regarding the presence and frequencies of mutations/polymorphisms in both the coding and non-coding regions of the mitochondrial genome, as well as data reporting mtDNA copy numbers, was extracted from studies. Information regarding authorship, geographical location, and sample size was also extracted.

In cases where data was only available in graphical form, Automeris WebPlotDigitizer, a web-based software tool designed specifically for graph reading, was employed to improve the accuracy of the values extracted.

In order to optimise the completeness of data, study authors were contacted regarding missing datasets, and supplementary data sources were searched where possible.

### 3.4 Data Synthesis and Statistical Analysis

All data was managed by the author, with professional advice sought from an independent statistician. Of the 15 studies included, 8 had sufficient data to be considered for various meta-analyses. Forest plots and graphical representation of figures were carried out using Review-Manager 5.4.1 (RevMan) statistical analysis software. Heterogeneity between studies was quantified by Cochran’s-Q scores and inconsistency square (I^2^) measures, with values of I^2^>50% indicating significant heterogeneity.

Quantitative data regarding genetic variants was processed to represent data as “percentage frequency of a given polymorphism (%)” in both PCOS and control groups. Where possible, this data was shown in forest plots, and reported as odds ratios (OR), along with respective 95% CIs, p-values, and heterogeneity (I^2^) scores.

Data regarding mtDNA content was processed to represent data as “log-transformed mean mtDNA copy number” (log [mean mtDNA CN]), and studies showing suitable measures of spread, such as standard deviation (SD) or standard error of the mean (SEM), were used to construct a forest plot. Subgroup analyses were performed to consider separately studies which assessed mtDNA copy number in blood cells, compared to 1 study which assessed mtDNA copy number in granulosa cells. A pooled estimate of the standardised mean difference (SMD) in log-transformed mean mtDNA copy number was calculated, again with a 95% CI, p-value, and heterogeneity (I^2^) score.

Where quantitative measures were not feasible, data was reported and analysed in a qualitative manner. Despite original intentions to, funnel plots could not be constructed to assess risk of publication biases, owing to the limited numbers of studies considering each outcome.

### 3.5 Quality Assessment

The quality of all included studies was assessed using the Q-Genie Tool for Quality Assessment of-Genetic Association Studies,[12,13] considering the following 11 categories:

1. Rationale for study
2. Selection and definition of outcomes of interest
3. Selection and comparability of comparison groups
4. Technical classification of exposure
5. Non-technical classification of exposure
6. Sources of bias
7. Sample size and power
8. A priori planning of analyses
9. Statistical methods and control for confounding
10. Testing of assumptions and inferences for genetic analyses
11. Appropriateness of inferences drawn from results

Studies were awarded scores between 1 (poor) and 7 (excellent) for each category. Total quality scores were calculated for each study, with scores ≤35 indicating poor quality, scores >35 and ≤45 indicating moderate quality, and scores >45 indicating good quality. Median scores and interquartile ranges (IQR) for all 11 categories were also calculated.

## 4. RESULTS

### 4.1 Search Results

The overall search results are summarised in ***Figure 1***. 13,812 records were identified through searching, spanning years 1964-2021. Following the removal of 3,767 duplicates, 10,045 records were screened for eligibility, first by title, excluding 5,938 records, then by abstract, excluding a further 4,026 records. After considering the remaining 81 full-text records, a further 67 records were excluded, thus leaving 14 studies eligible for inclusion. After searching reference lists, 1 additional study was added. Overall, this systematic review included 15 studies (shown in ***Table 1***), considering a total of 2,053 participants (1,125 PCOS, 928 control). Of these studies, 8 had suitable data to be included in the meta-analysis.

**Table 1.** Studies included in systematic review.

|  | Study ID | Title of Publication | Location | Available at |
| --- | --- | --- | --- | --- |
| INCLUDED IN META-ANALYSIS | **Hu et al. (2011)[14]** | Single Nucleotide Polymorphisms (SNPs) And Variable Number Tandem Repeats (VNTRs) In MtDNA D-Loop and CO II- tRNA Lys Intergenic Region With PCOS | China | http://dx.doi.org/10.1016/S1001-7844%2812%2960008-X |
|  | **Zhuo et al. (2012)[15]** | Analysis of Mitochondrial DNA Sequence Variants in Patients with Polycystic Ovary Syndrome | China | https://dx.doi.org/10.1007/s00404-012-2358-7 |
|  | **Ding et al. (2017)[18]** | Mutations in Mitochondrial tRNA Genes May Be Related to Insulin Resistance in Women with Polycystic Ovary Syndrome | China | http://www.ajtr.org/files/ajtr0048638.pdf |
|  | **Reddy et al. (2019)[6]** | Impact of Mitochondrial DNA Copy Number and Displacement Loop Alterations on Polycystic Ovary Syndrome Risk in South Indian Women | India | https://dx.doi.org/10.1016/j.mito.2017.12.010 |
|  | **Saeed et al. (2019)[20]** | Polycystic Ovary Syndrome Dependency on MtDNA Mutation; Copy Number and Its Association with Insulin Resistance | Iraq | https://dx.doi.org/10.1186/s13104-019-4453-3 |
|  | **Shukla et al. (2020)[22]** | Identification of Variants in Mitochondrial D-Loop and OriL Region and Analysis of Mitochondrial DNA Copy Number in Women with Polycystic Ovary Syndrome | India | https://dx.doi.org/10.1089/dna.2019.5323 |
| | **Wang et al. (2020)[23]** | The Effects of Mitochondrial Dysfunction on Energy Metabolism Switch By HIF-1α Signaling in Granulosa Cells of Polycystic Ovary Syndrome | China | http://dx.doi.org/10.5603/EP.a2020.0002 |
|  | **Deng et al. (2021)[33]** | Polymorphisms and Haplotype of Mitochondrial DNA D-Loop Region Are Associated with Polycystic Ovary Syndrome in A Chinese Population | China | http://dx.doi.org/10.1016/j.mito.2020.12.006 |
| INSUFFICIENT DATA FOR META-ANALYSIS | **Zhuo et al. (2010)[16]** | A 9-Bp Deletion Homoplasmy In Women with Polycystic Ovary Syndrome Revealed by Mitochondrial Genome-Mutation Screen | China | https://dx.doi.org/10.1007/s10528-009-9308-5 |
|  | **Lee et al. (2011)[43]** | Mitochondrial DNA Copy Number in Peripheral Blood in Polycystic Ovary Syndrome | Korea | https://dx.doi.org/10.1016/j.metabol.2011.04.010 |
|  | **Rabøl et al. (2011)[49]** | Skeletal Muscle Mitochondrial Function in Polycystic Ovarian Syndrome | Denmark | https://dx.doi.org/10.1530/EJE-11-0419 |
|  | **Ding et al. (2016a)[21]** | The Mitochondrial tRNA<sup>Leu</sup> (UUR) A3302G Mutation May Be Associated with Insulin Resistance in Women with Polycystic Ovary Syndrome | China | https://dx.doi.org/10.1177/1933719115602777 |
|  | **Ding et al. (2016b)[19]** | Point Mutation in Mitochondrial tRNA Gene Is Associated with Polycystic Ovary Syndrome and Insulin Resistance | China | https://dx.doi.org/10.3892/mmr.2016.4916 |
|  | **Ding et al. (2018)[17]** | Mitochondrial tRNA<sup>Leu</sup> (UUR) C3275T, tRNA<sup>Gln</sup> T4363C And tRNA<sup>Lys</sup> A8343G Mutations May Be Associated with PCOS And Metabolic Syndrome | China | https://dx.doi.org/10.1016/j.gene.2017.11.049 |
|  | **Yang et al. (2020)[50]** | Changes in Peripheral Mitochondrial DNA Copy Number in Women with Polycystic Ovary Syndrome: A Longitudinal Study | Taiwan | http://dx.doi.org/10.1186/s12958-020-00629-5 |

**Figure 1.**
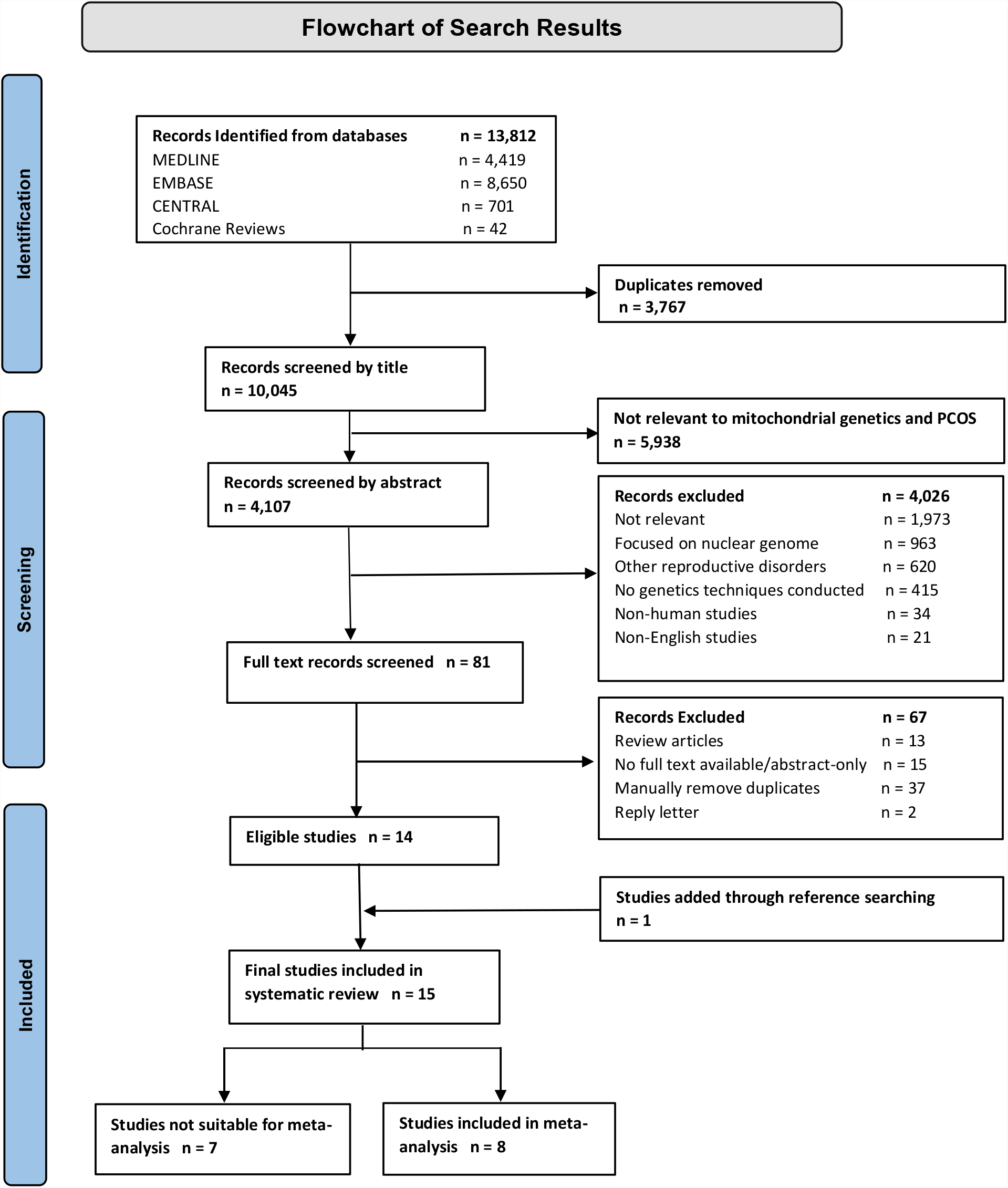
Flowchart of search results and included studies

### 4.2 Genetic Variants in Coding Regions of the Mitochondrial Genome

Polymorphisms in the coding region of the mitochondrial genome were reported by 7 studies.

#### 4.2.1 9-Base Pair Deletions

Four studies documented the presence of a 9-bp deletion (CCCCCTCTA) in region V of the mitochondrial genome.[14-17] Whilst all 4 studies agreed that this deletion occurred more frequently in women with PCOS than in controls, only 1 study quantified this effect, showing this deletion to occur in 23.5% of cases compared to 7.1% of controls, and reported a p-value of p=0.02, indicating a significant relationship.[16]

#### 4.2.2 Polymorphisms in the ND5 and A6 Genes

The full list of all the genetic polymorphisms detected in other coding regions is available in ***Supplement 2***.

Frequently reported variants included the T12811C and T12338C polymorphisms in the ND5 gene,[15,18,19] and the G8584A and C8684T polymorphisms in the A6 gene.[15,16,18] Whilst sufficient numerical data was not available to calculate the frequencies of these polymorphisms, these mutations were detected only in PCOS cases, and not in controls, eluding to their potential significance.

#### 4.2.3 Polymorphisms in Genes Coding for Mitochondrial tRNAs and rRNAs

All data extracted considering mt-tRNA genes is available in ***Supplement 3***.

Polymorphisms appeared to occur most frequently in genes coding for the following 7 mt-tRNAs: tRNA^Cys^, tRNA^Leu^, tRNA^Glu^, tRNA^Gln^, tRNA^Lys^, tRNA^Arg^, and tRNA^Asp^. The 3 studies with sufficient data were used to determine the frequency of polymorphisms in these tRNAs which are summarised in ***Figure 2a***.[15,18,20] tRNA^Cys^ gene polymorphisms were more frequent in PCOS cases than in controls, occurring in 2.9% of-cases vs. 1.1% of controls. This was also the case for polymorphisms in genes coding for tRNA^Leu^, which occurred in 2.7% of cases, but only in 0.6% of controls. Polymorphisms in genes coding for tRNA^Glu^, tRNA^Gln^, tRNA^Lys^, tRNA^Arg^, and tRNA^Asp^, were detected only in PCOS cases and not in controls, occurring at frequencies of: 13.1%, 3.1%, 2.2%, 1.9%, and 1.5% respectively. Overall, mt-tRNA gene polymorphisms reported in PCOS cases were either absent, or less frequent, in controls.

**Figure 2.**
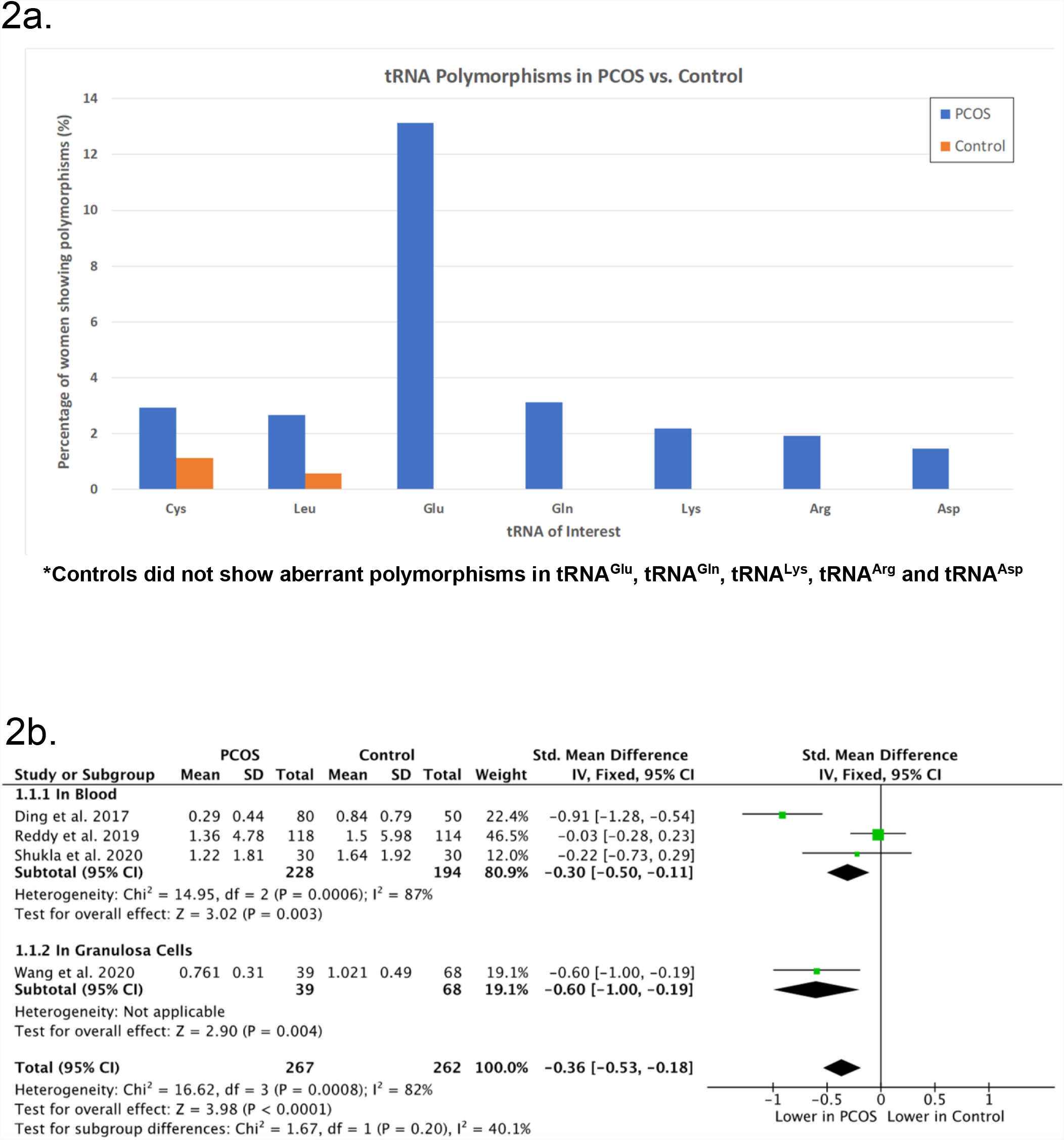
a) Bar graph of mitochondrial tRNA gene polymorphisms b) Log-transformed mean mtDNA copy number in PCOS cases vs. controls

Polymorphisms in genes coding for both the 12S and 16S rRNAs were also detected in women with PCOS (***Supplement 4***).[15-17,21]

### 4.3 Genetic Variants in Non-Coding Regions of the Mitochondrial Genome

#### 4.3.1 Polymorphisms in the Mitochondrial D-Loop

D-Loop polymorphisms were considered by 8 studies, with all extractable data shown in ***Supplement 5***. The following 5 polymorphisms were reported in 2 or more studies: C150T, T146C, A263G, A189G, D310. However, only 3 of these polymorphisms, C150T, T146C, and C150T, had sufficient data available to be considered for meta-analysis (***Figure 3)***.

**Figure 3.**
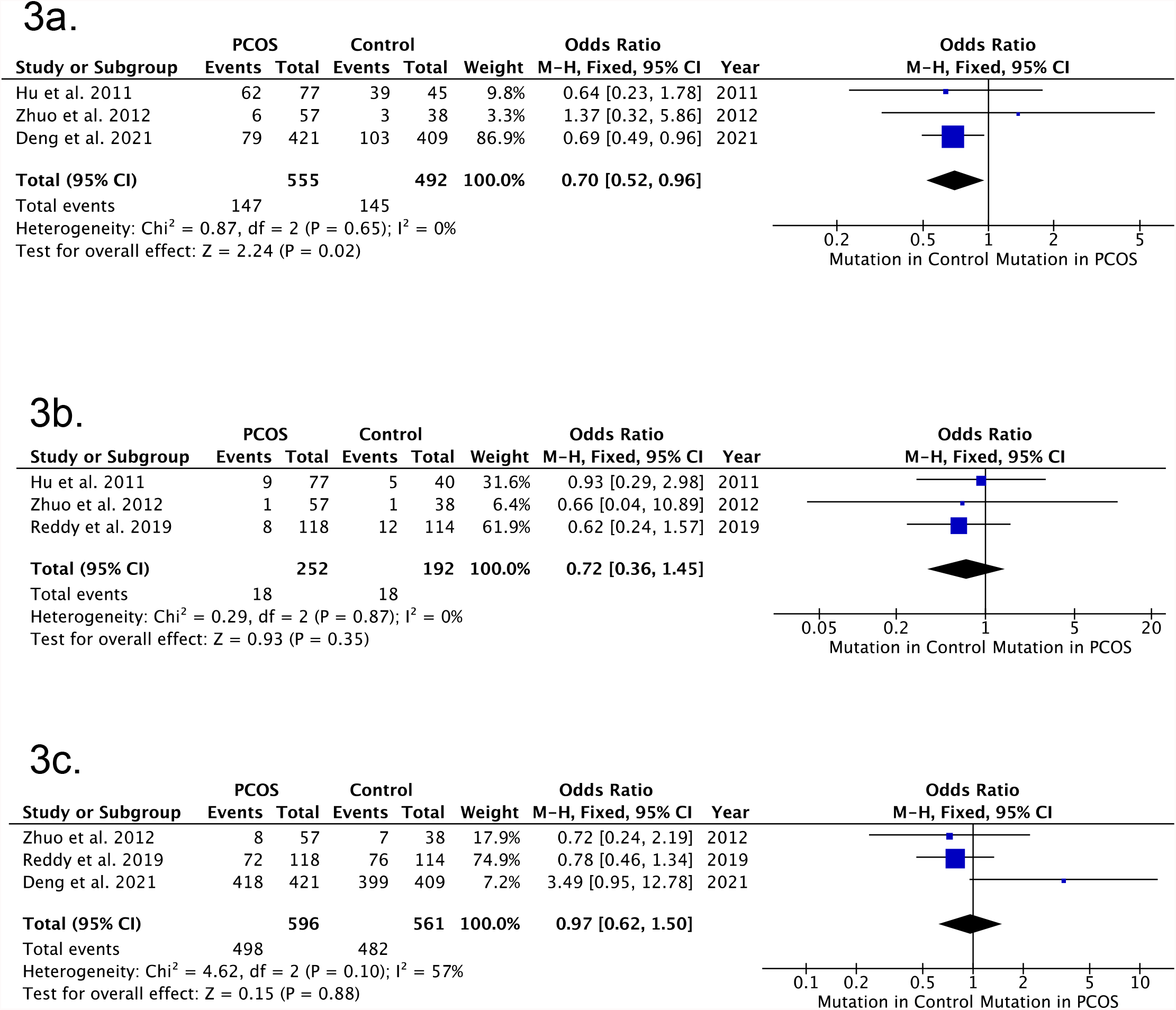
a) C150T D-Loop polymorphisms in PCOS cases vs. controls b) T146C D-Loop polymorphisms in PCOS cases vs. controls c) A263G D-Loop polymorphisms in PCOS cases vs. controls

As shown in ***Figure 3a***, the C150T D-Loop polymorphism occurred at a significantly lower frequency in PCOS cases compared to controls, with an overall odds ratio (OR) of 0.70 (95% CI 0.52, 0.96, p=0.02). Whilst the T146C polymorphism (***Figure 3b***) also occurred at a lower frequency in PCOS cases than in controls, with an overall OR of 0.72 (95% CI-0.36, 1.45, p=0.35), this result is not considered to be significant. As shown by the OR of 0.97 (95%CI 0.62, 1.50, p=0.88), the A263G polymorphism (***Figure 3c***) in the D-Loop of the mitochondrial genome occurred at similar frequencies in both cases and controls, again indicating a result that is not significant.

Overall, women with PCOS showed lower odds of possessing the C150T polymorphism, potentially lower odds of possessing the T146C polymorphism, and almost the same odds of possessing the A263G polymorphism, as controls.

One study also detected the A189G and D310 polymorphisms almost twice as often in women with PCOS compared to controls, with the A189G polymorphism present in 26.3% of PCOS cases vs. 13.2% of controls, and the D130 polymorphism present in 54.2% of-cases vs. 28.9% of-controls.[6] Interestingly, women with PCOS who showed these two specific polymorphisms appeared to have significantly lower mtDNA-copy numbers than those who did not show these polymorphisms.

### 4.4 Changes in Mitochondrial Genetic Content

Mitochondrial genetic content, reported as log-transformed mtDNA copy number, was assessed by 9 studies (***Supplement 6***).

Whilst all 9 studies showed lowered mtDNA copy numbers in women with PCOS, 1 study did not report data for the control group, and 4 did not report suitable measures of spread to be considered for meta-analysis. The data from the 4 remaining studies,[6,18,22,23] considering a total of 529 subjects (267 PCOS, 262 control), is summarised in ***Figure 2b***.

Subgroup analyses were adopted to consider separately 3 studies which assessed mtDNA copy number in blood leukocytes, compared to 1 study which assessed mtDNA copy number in granulosa cells.

Log-transformed mean mtDNA copy number was significantly reduced in PCOS cases compared to controls, with an overall standardised mean difference (SMD) of--0.36 (95% CI-0.53, -0.18, p<0.0001). Considering the subgroup analyses, log-transformed mean copy number was shown to be significantly reduced in both blood and granulosa cells with SMD values of -0.30 (95%-CI, -0.50, -0.11) and -0.60 (95%-CI, -1.00, -0.19) respectively. The value of I^2^=82% does, however, indicate significant between-study heterogeneity.

### 4.5 Quality assessment

Quality assessment was carried out for all included studies, with each study being assigned an overall quality score (***Supplement 7***). The majority of papers (6/15, 40%) were found to be of moderate quality, whilst the minority of papers (4/15, 26.7%) were deemed to be of poor quality. An intermediate number of papers (5/15, 33.3%) were regarded as good quality.

The highest scoring category was “Rationale for Study” (median 6, IQR 4-7), whilst the two poorest performing categories included “Other Sources of Bias” (median 2, IQR 1-3), and “Sample Size and Power” (median 2, IQR 1-2).

## 5. DISCUSSION

A current gap in literature exists relating to a systematic review of the associations between mitochondrial genetics and PCOS. As such, this study aimed to assess genetic variants in both the coding and non-coding regions of the mitochondrial genome, and changes in mitochondrial genetic content in women with PCOS, whilst evaluating the quality of evidence overall.

### 5.1 Polymorphisms in Coding Regions of the Mitochondrial Genome

In the majority of controls without PCOS, the 9-bp sequence (CCCCCTCTA) in region V of the mitochondrial genome was repeated twice in tandem. Many women with PCOS, however, showed deletions in this region, leaving them with only one copy of this sequence. These deletions are suspected to have arisen from DNA replication slippage errors made by DNA polymerase-gamma, which possesses underdeveloped repair mechanisms in mtDNA.[16,24] Interestingly, Hu et al. found these deletions to be significantly associated with higher serum levels of glucose during oral glucose tolerance testing (OGTT), and lower insulin sensitivity indices,[14] suggesting that the deletions may be an underlying factor contributing towards insulin resistant states in PCOS patients.

The ND5 T12811C and T12338C polymorphisms, as well as the A6 G8584A and C8684T polymorphisms, were detected only in PCOS cases, and not in controls. These polymorphisms were found to cause amino acid substitutions,[15,19] and could therefore bear important implications. Amino acid changes result in altered primary, and potentially secondary/tertiary/quaternary structures of proteins. This could subsequently impact the ETC, which itself is a series of proteins, and could then lead to inefficiencies in electron transfer and higher levels of ROS. These genetic variants, given that they were only found in cases, may therefore contribute to the wide-spread metabolic dysfunction witnessed in PCOS.

Interestingly, all four SNPs listed above have been associated with Leber’s Hereditary Optic Neuropathy (LHON) Syndrome,[25-27] with the ND5 T12338C mutation also being linked to neonatal deafness.[28] Both of these disorders showcase failures in energy metabolism, and are known to be mitochondrially, and therefore maternally, inherited. This may suggest that some characteristics of PCOS, having shown such similarities in mitochondrial genetics and energy disruptions (albeit to a lesser extent), may also be attributed to a more maternal pattern of inheritance, by way of the mitochondrial genome.

### 5.2 Genes Coding for tRNA AND rRNA

Polymorphisms in genes coding for tRNAs and rRNAs were present and much more frequent in PCOS cases compared to controls. This too could bear consequences for women with PCOS, considering the essential roles of tRNAs and rRNAs in the translation of mitochondrial-encoded proteins. The majority of detected variants occurred in regions that remain highly conserved amongst vertebrate species, eluding to their ability to alter pivotal mitochondrial processes, and to result in biochemical changes and oxidative stress, as seen in PCOS.

Some polymorphisms reported in women with PCOS, including the tRNA^Gln^ T4395C and tRNA^Leu^ A3302G polymorphisms,[15,21] occurred in the acceptor arms of tRNA molecules -the regions which attach to amino acids in the process of aminoacylation. Such genetic variants may therefore possess the ability to disrupt amino acid-tRNAs binding, subsequently impacting the assembly of polypeptide chains. Other polymorphisms, such as the tRNA^Asp^ A7543G, and tRNA^Leu^ T4363C polymorphisms, occurred in the anticodon stem of tRNA molecules,[18] and could potentially disrupt base-pairing with mRNA transcripts,[18] and cause inefficiencies in the link between transcription and translation of mitochondrial-encoded proteins. Other SNPs, including the tRNA^Lys^ A8343G, tRNA^Arg^ T10454C, and tRNA^Glu^ A14693G SNPs,[18] were localised to the T-Loop of tRNA molecules, which is an important recognition site that allows tRNA molecules to bind to mitochondrial ribosomes, which themselves appeared to bear genetic polymorphisms in women with PCOS.

Current data suggests that these polymorphisms confer structural and functional changes in the mitochondrial protein synthesising machinery, and result in destabilisation of tRNA and rRNA molecules,[29] loss of nucleotide modifications, and deficient aminoacylation,[30,31] all of which may affect tRNA-amino acid binding, and tRNA-ribosome complex formation.[32] Such alterations in protein synthesis may once again affect complexes that make up the ETC, and may offer possible explanations for the altered enzyme levels seen in women with PCOS.[30] As evidence of this, women carrying these mutations were found to have lower levels of superoxide dismutase, a key enzyme and hence protein, as well as higher levels of oxidative stress markers, such as 8-hydroxy-2′–deoxyguanosine (8-OHdG) and malondialdehyde (MDA).[18] Women with these tRNA mutations also showed lower mtDNA copy numbers, eluding to disturbances in mitochondrial equilibrium.[18]

Mt-tRNA gene polymorphisms also demonstrated links to other mitochondrially-inherited diseases, with similar biochemical and metabolic profiles to those of PCOS. In a familial study, Ding et al. suggested that the tRNA^Leu^ C3275T, tRNA^Gln^ T4363C, and tRNA^Lys^ A8343G SNPs were associated with maternally-inherited MetS,[17] whilst in another familial study it was suggested that the tRNA^Leu^ A3302G SNP was associated with maternally-inherited IR.[21] This evidence further supports that the pathology presented in PCOS may be partly rooted in mitochondrial genetics.

### 5.3 Polymorphisms in the Non-Coding Region of the Mitochondrial Genome

Unlike the above polymorphisms, the D-Loop C150T polymorphism and, more often than not, the T146C polymorphism, occurred at higher frequencies in controls, suggesting that these polymorphisms may have a somewhat protective element that contributes a slightly lowered risk of PCOS.[33] Interestingly, the T146C polymorphism has also been associated with lowered risk of endometriosis,[34,35] indicating a potentially key polymorphism in wider female reproduction. In contrast, the A263G polymorphism occurred at similar frequencies in both cases and controls, with some studies indicating a significant relationship and others not, summating to show no overall effect. Inconsistencies between studies could have arisen due to variations in the prevalence of this SNP, owing to ethnic or environmental differences.

Whilst genetic variants in the D-Loop do not affect mitochondrial-encoded proteins directly, they have the ability to alter gene expression regulation, and to bring about changes in mtDNA replication and transcription, given that the D-Loop contains the origins of replication of the mitochondrial genome.[36] As such, genetic polymorphisms in the D-loop have the capacity to influence mitochondrial dynamics, and may therefore contribute further to ROS production in PCOS.

As an example of this, women with PCOS carrying the D-Loop A189G and D310 polymorphisms were shown to have significantly lower mtDNA copy numbers than those not carrying these polymorphisms.[6] The D310 region is located within one of the key conserved regions of the genome (CSB-II) essential for mtDNA replication,[6,37] offering a potential explanation as to why PCOS cases with D310 variants show lowered mtDNA copy numbers, and subsequently experience oxidative stress.

Apart from regulating the mitochondrial genome, the D-loop also enables nuclear-mitochondrial crosstalk. Multiple nuclear hormone receptors, including the glucocorticoid receptors, thyroid receptors (T3), and oestrogen receptors, bind to regions of the D-Loop as part of their normal physiological function,[22,38] creating scope for genetic variants in these regions to influence hormone profiles in women with PCOS, and the endocrine system at large.

### 5.4 Mitochondrial DNA Content

Compared to controls, PCOS cases showed significantly lowered mtDNA copy numbers, indicating a reduction in both mitochondrial number and mass overall. This is likely to be a result of autophagy,[39] a process by which cells degrade their damaged organelles, including mitochondria, by lysosomal action.[40] This decrease may be testament to the effects of harbouring these various genetic variants, which act to increase ROS production to the point of cell death. The reduction of mitochondrial content in both blood and granulosa cells suggests that this mitochondrial dysfunction not only affects cells of the ovary, but also bears more widespread consequences in women with PCOS, causing this syndrome to present with multi-organ and multi-system involvement.

Studies have shown that maintenance of mtDNA copy numbers is essential for preservation of mitochondrial function and cell growth.[41] A decrease in mitochondrial content translates to a subsequent decrease in the expression of mitochondrial-encoded genes and proteins. This means that a lowered number of mitochondria and mtDNA-encoded enzymes are left to manage the raised level of ROS after damaged mitochondria are removed, predisposing the remaining mitochondria to further damage, oxidative stress, and inevitable autophagy. Therefore, a diminished mtDNA copy number in PCOS may not only be a result of increased ROS generation, but may contribute further towards it, by feeding into the “vicious cycle” of ROS-induced ROS release.[42]

PCOS patients with lowered mtDNA copy numbers have also shown to have higher degrees of IR, as well as lowered levels of sex hormone binding globulin (SHBG),[43] suggesting that mtDNA depletion could be involved in the pathogenesis of this syndrome, linking changes in genetic expression to hormonal and homeostatic imbalances.

These findings are further supported by multiple animal studies, which showed abnormalities in mitochondrial structure and function, and reported alterations in expression of mtDNA-encoded genes as a result of lower mtDNA copy numbers.[44,45] Reduced mtDNA content has also been linked to poor oocyte quality,[46] and as such, could contribute to subfertility or infertility in women with PCOS.

### 5.5 The Mitochondrial Genome in Women with PCOS

Overall, women with PCOS showcased polymorphisms in both coding and non-coding regions of the mitochondrial genome, and showed a reduction in the amount of mtDNA present. These genetic variants have the ability to affect mitochondrial function on multiple levels, be it at the level of replication and transcription through D-Loop polymorphisms or slippage errors, or at the level of translation, by way of polymorphisms in genes coding for proteins, tRNAs, and rRNAs.

Based on the above findings, this study proposes that this “multi-level” genetic dysregulation sets up a cycle of events that then predisposes to further genetic damage and mitochondrial destruction, with the cycle unfolding as follows: initial genetic variants contribute to impairment of ETC function,[30] which subsequently leads to less efficient electron transfer, and therefore increased rates of ROS generation, bringing about oxidative stress. This oxidative stress then burdens cells to the point of apoptosis,[39,40] resulting in lowered mitochondrial content, which subsequently leaves a reduced pool of mitochondria in an environment of increased oxidative stress, thus leading to further mtDNA damage (***Figure 4***). This proposed cycle draws parallels with the concept of ROS-induced ROS release,[42] but additionally accounts for the presence of aberrant genetic variants in the mitochondrial genomes of women with PCOS.

**Figure 4.**
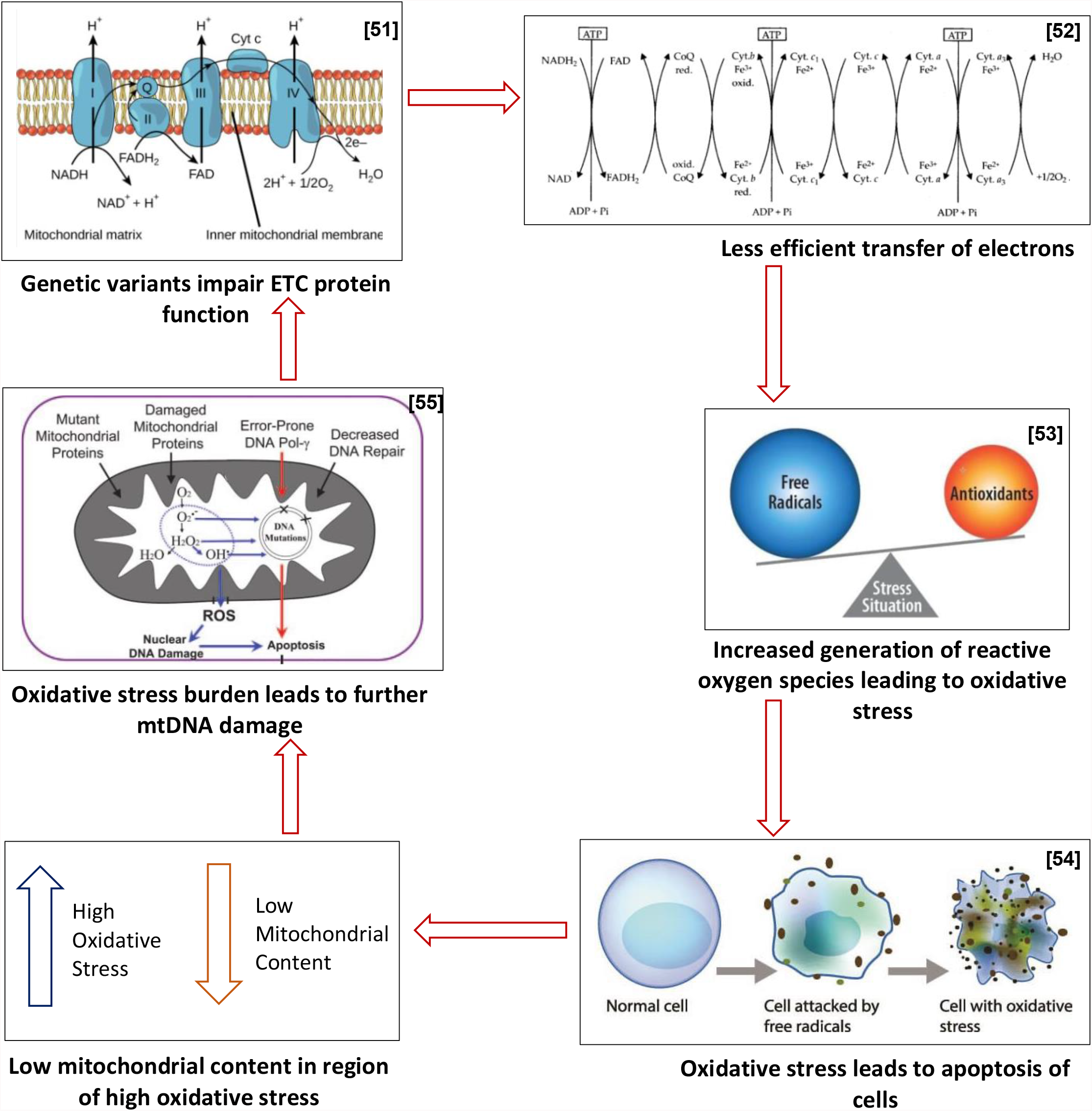
Proposed cycle of events in women with PCOS

### 5.6 Quality Assessment and Critical Appraisal of Studies

Studies scored highly in the category entitled “Rationale for study”, which subsequently translated to the majority of studies possessing clear hypotheses. Studies also benefitted from having well-defined eligibility criteria for cases, with all but two studies characterising PCOS cases as per the revised 2003 Rotterdam criteria.[47] Studies also showcased well-structured methodology, and use of accredited statistical software, showing good *a priori* planning of analyses. Studies achieving the highest scores were those which documented better genetic techniques, for example those which detailed efforts to normalise expression of target genes relative to housekeeping genes (B-globin/ B-actin/ GAPDH).

However, studies appeared to be insufficiently powered by generally small sample sizes, and did not appropriately account for possible sources of bias. Many studies recruited cases from hospitals or fertility clinics. Therefore, in order to be considered as cases, women would have had to experience subfertility/infertility, or symptoms considerable enough to seek professional intervention, arguably representing only the most severe cases of PCOS. By only considering women at one end of the spectrum of what is a largely heterogenous disorder, studies were made vulnerable to selection bias, as cases sampled may not be representative of wider PCOS populations.

Similarly, whilst studies recruited age-matched and BMI-matched controls, other important characteristics, such as smoking– a factor known to cause genetic mutagenesis, and diet – a factor important in insulin resistant PCOS states, were less regulated, potentially introducing heterogeneity between participants.

A minority of studies also failed to show records of all collected data raising concerns around publication biases. Owing to the limited number of papers, however, the use of funnel plots to assess risks of publication biases was deemed inappropriate for this study.

Moreover, only 8 studies were fit for inclusion in various meta-analyses, with other studies showcasing insufficient data to undergo appropriate statistical analysis. Some studies successfully reported mtDNA copy numbers, but failed to report suitable measures of spread to be included in forest plots. Other studies documented an array of genetic variants, but failed to report numerical data that allowed for the frequencies of polymorphisms to be calculated. As such, much of the extracted data either could not be analysed to its full potential, or was limited by its qualitative nature. Whilst the outcomes of such papers were still assessed qualitatively, these findings could not be quantified in a manner that allowed for statistical validation of results.

Overall, only 5/15 (33.3%) of studies were found to be of good quality. This has potential implications on the outcomes of this review, as it becomes difficult to gauge the true relationship between the mitochondrial genome and PCOS.

### 5.7 Strengths and Limitations

To the authors’ knowledge, this is the first systematic review to explore the relationship between the mitochondrial genome and PCOS. Not only does this study address a gap in previous literature, but it also focuses on a newly emerging concept, making it significant to on-going and future research. However, by restricting the search to English-only results, and by only accounting for literature published on online platform, some otherwise relevant studies may have been excluded from this review. By its design, this review was also based on the assumption that data presented in primary studies was valid and accurate. Moreover, this review, much like its included studies, considered genetic variants in the mitochondrial genomes of women who had already obtained a diagnosis of PCOS, making it difficult to distinguish cause from effect, and to gauge the true association between the mitochondrial genome and PCOS.

### 5.8 Future Implications

In clinical practice, women with PCOS are treated with Metformin to encourage normalised glucose usage in cells.[48] This study has shown that if this regulation is not achieved, blood leukocytes will experience oxidative stress and undergo apoptosis, leading to reduced mtDNA copy number. In theory, this could mean that mtDNA copy numbers measured in blood samples could be used as a minimally invasive way to monitor Metformin treatment and efficacy in women with PCOS, with lower mtDNA copy number indicating higher rates of cell death and less effective treatment. Women are also commonly advised to incorporate foods with lower glycaemic indices (GI) into their diets. Given the capacity of oxidative stress to disrupt mitochondrial equilibrium and genetics, women with PCOS may also be advised to increase intake of foods rich in anti-oxidants.

### 5.9 Future Recommendations

Due to the aforementioned data limitations, this study proposes the implementation of a standardised protocol for reporting in genetic association studies. Abiding by such a protocol will allow uniform reporting of data that can then be processed to quantify the relationship between the mitochondrial genome and PCOS more accurately.

Future research should also aim to explore different aspects of the mitochondria in the context of PCOS. Whilst many studies considered mitochondrial content in blood leukocytes, only 2 were found to consider mitochondrial content in granulosa cells, with only one being suitable for meta-analysis. To allow for larger subgroup analyses, and to uncover a more holistic relationship, it is recommended that more studies investigate mitochondrial genomes in granulosa cells, and in other cell types involved PCOS, such as beta-pancreatic cells given their role in insulin disturbances. It is also worth noting that the majority of the studies were conducted in Asian populations, and it is necessary to explore this relationship in women of other ethnicities and genetic backgrounds.

More familial studies, carrying out genetic screening of maternal mitochondrial genomes, could help decipher whether mitochondrial genetic variants are germline or acquired changes, and may help determine a cause-effect relationship.

Moreover, the mitochondrial genome, although distinct from the nuclear genome, does not function independently of nuclear-encoded proteins. Future research should therefore aim to explore mitochondrial genetics in women with PCOS in a more expansive manner to account for nuclear-mitochondrial crosstalk.

These recommendations would provide valuable insight on the topic of mitochondrial genetics and PCOS, and would allow for this relationship to be put into context with respect to wider bodily function and inheritance patterns.

## 6. CONCLUSION

Women with PCOS appear to harbour genetic variants in both coding and non-coding regions of their mitochondrial genomes, including 9-bp deletions and an array of specific SNPs. These are thought to contribute to aberrant protein synthesis of mitochondrial elements, and to result in oxidative stress in cells, leading to autophagy, and thus, lowered mitochondrial content. This is likely to feed into a cycle of ROS-induced ROS release, predisposing to further mitochondrial dysfunction.

Given that only 33.3% of analysed studies were found to be of good quality, however, further research, reporting more quantitative data regarding different cell types and women of different ethnicities, is required to unveil to the true association between mitochondrial genetics and PCOS, and to understand future management of this complex syndrome.

## Data Availability

All data produced in the present work are contained in the manuscript

## 7. ACKNOWLEDGMENTS

Special thanks to Dr Aviva Petrie, for her professional advice regarding data management and statistical analyses, and for her educational support.

Further thanks to the authors of the various studies included in this systematic review, for providing the data and framework on which this study was based.

