## Supplements 1-7 for "Genetic Associations with Polycystic Ovary Syndrome: The Role of The Mitochondrial Genome; A Systematic Review and Meta-analysis"

#### SUPPLEMENT 1 – SEARCH TERMS

**Search terms relating to "Mitochondrial Genetics"**

Association analysis
Candidate gene
Circular DNA
Coding region
Copy number
Copy number variation
D-loop
Deep sequencing
Deletion
Displacement loop
DNA
DNA polymerase-gamma
DNA sequence
Electron transport chain
ETC
Gen* linkage
Gen* mapping
Gen* mutation
Gene expression
Gene expression profiling
Gene expression regulation
Gene frequency
Gene* profile
Gene regulatory network
Gene sequencing
Gene* variant
Genes
Genetic analysis
Genetic association
Genetic association study
Genetic background
Genetic defect
Genetic epidemiology
Genetic factor
Genetic influence
Genetic linkage
Genetic marker
Genetic polymorphism
Genetic predisposition to disease
Genetic stud*
Genetic susceptibility
Genetic techniqu*
Genetic test*
Genetic variation
Genetic*
Genome
Genome analysis
Genome-wide
Genome-wide association
Genomics
Haplotype
Heavy strand
HVR-1
HVR-2
Hypervariable
Hypervariable region
Hypervariable region-1
Hypervariable region-2
Insertion
Light strand
Messenger RNA
Microsatellite repeat
Mitochondrial DNA
Mitochondrial genome
Mitochondrial-encoded
Mitoribosome
mtDNA
Multifactorial inheritance
Mutation
Next generation sequencing
NGS
Non-coding region
Nucleotide sequencing
Organelle
Oxidative stress
PCR
Phenotype
Polygenic
Polymerase chain reaction
Polymorphism
Promoter region
Reactive oxygen species
Respiratory chain
Respiratory chain complex
ROS
Single nucleotide polymorphism
SNP
Southern blot
Tandem repeat
Transcript*
Transcription factor
Translation
Trinucleotide repeat
Western blot
Whole exome sequencing

***Search terms relating to PCOS***

Multicystic ovary
Polycystic ovary
PCO
PCOS
Polycystic disease of the ovary
Polycystic Ovarian Disease
Polycystic Ovarian Syndrome
Polycystic ovaries
Polycystic ovary disease
Polycystic ovary morphology
Polycystic Ovary Syndrome
POS
Stein-Leventhal Syndrome
Stein Leventhal Syndrome

### SUPPLEMENT 2 – POLYMORPHISMS IN CODING REGIONS OF THE MITOCHONDRIAL GENOME

| Study ID | Gene | Position |
|---|---|---|
| Zhuo 2010 | A6 | 8584 |
| Zhuo 2010 | A6 | 8701 |
| Zhuo 2010 | A6 | 8745 |
| Zhuo 2010 | A6 | 8794 |
| Zhuo 2010 | A6 | 9053 |
| Zhuo 2010 | A6 | 9101 |
| Zhuo 2010 | A6 | 9128 |
| Zhuo 2010 | A6 | 9180 |
| Zhuo 2012 | A6 | 8584 |
| Zhuo 2012 | A6 | 8684 |
| Zhuo 2012 | A6 | 8745 |
| Zhuo 2012 | A6 | 8793 |
| Zhuo 2012 | A6 | 8860 |
| Zhuo 2012 | A6 | 9053 |
| Zhuo 2012 | A6 | 9126 |
| Zhuo 2012 | A6 | 9128 |
| Ding 2016 | A6 | 8585 |
| Ding 2016 | A6 | 8860 |
| Ding 2017 | A6 | 8584 |
| Ding 2017 | A6 | 8684 |
| Zhuo 2010 | A8 | 8392 |
| Zhuo 2010 | A8 | 8414 |
| Zhuo 2010 | A8 | 8459 |
| Zhuo 2010 | A8 | 8473 |
| Zhuo 2012 | A8 | 8392 |
| Zhuo 2012 | A8 | 8414 |
| Zhuo 2012 | A8 | 8459 |
| Zhuo 2012 | A8 | 8473 |
| Ding 2016 | A8 | 8414 |
| Zhuo 2010 | COX1 | 6392 |
| Zhuo 2010 | COX1 | 6962 |
| Ding 2016 | COX1 | 6392 |
| Ding 2016 | COX1 | 7028 |
| Ding 2018 | COX1 | 7028 |
| Zhuo 2012 | COX2 | 6221 |
| Zhuo 2012 | COX2 | 6272 |
| Zhuo 2012 | COX2 | 6338 |
| Zhuo 2012 | COX2 | 6392 |
| Zhuo 2012 | COX2 | 6455 |
| Zhuo 2012 | COX2 | 6962 |
| Zhuo 2012 | COX2 | 7028 |
| Zhuo 2012 | COX2 | 7142 |
| Zhuo 2010 | COX2 | 7853 |
| Ding 2016 | COX2 | 8020 |

| Study ID | Gene | Position |
|---|---|---|
| Zhuo 2010 | COX3 | 9536 |
| Zhuo 2010 | COX3 | 9540 |
| Zhuo 2010 | COX3 | 9548 |
| Zhuo 2010 | COX3 | 9824 |
| Zhuo 2012 | COX3 | 9242 |
| Zhuo 2012 | COX3 | 9336 |
| Zhuo 2012 | COX3 | 9377 |
| Zhuo 2012 | COX3 | 9540 |
| Zhuo 2012 | COX3 | 9559 |
| Zhuo 2012 | COX3 | 9824 |
| Ding 2016 | COX3 | 9540 |
| Ding 2016 | COX3 | 9824 |
| Zhuo 2010 | CYTB | 14783 |
| Zhuo 2010 | CYTB | 15038 |
| Zhuo 2010 | CYTB | 15043 |
| Zhuo 2010 | CYTB | 15244 |
| Zhuo 2010 | CYTB | 15346 |
| Zhuo 2010 | CYTB | 15301 |
| Zhuo 2012 | CYTB | 14783 |
| Zhuo 2012 | CYTB | 15043 |
| Zhuo 2012 | CYTB | 15244 |
| Zhuo 2012 | CYTB | 15301 |
| Zhuo 2012 | CYTB | 15326 |
| Zhuo 2012 | CYTB | 15460 |
| Zhuo 2012 | CYTB | 15724 |
| Ding 2016 | CYTB | 14766 |
| Ding 2016 | CYTB | 15301 |
| Ding 2016 | CYTB | 15326 |
| Ding 2018 | CYTB | 14766 |
| Ding 2018 | CYTB | 15326 |
| Ding 2018 | CYTB | 15535 |
| Zhuo 2010 | ND1 | 3316 |
| Zhuo 2010 | ND1 | 3394 |
| Zhuo 2010 | ND1 | 3497 |
| Zhuo 2010 | ND1 | 3970 |
| Zhuo 2010 | ND1 | 4086 |
| Zhuo 2010 | ND1 | 4248 |
| Zhuo 2012 | ND1 | 3316 |
| Zhuo 2012 | ND1 | 3394 |
| Zhuo 2012 | ND1 | 3423 |
| Zhuo 2012 | ND1 | 3497 |
| Zhuo 2012 | ND1 | 3552 |
| Zhuo 2012 | ND1 | 3834 |
| Zhuo 2012 | ND1 | 3970 |

| Study ID | Gene | Position |
|---|---|---|
| Zhuo 2012 | ND1 | 4086 |
| Ding 2016 | ND1 | 3553 |
| Ding 2016 | ND1 | 4047 |
| Ding 2017 | ND1 | 3394 |
| Zhuo 2010 | ND2 | 5147 |
| Zhuo 2010 | ND2 | 5178 |
| Zhuo 2010 | ND2 | 5301 |
| Zhuo 2010 | ND2 | 5417 |
| Zhuo 2010 | ND2 | 5460 |
| Zhuo 2012 | ND2 | 4688 |
| Zhuo 2012 | ND2 | 4769 |
| Zhuo 2012 | ND2 | 4883 |
| Zhuo 2012 | ND2 | 5046 |
| Zhuo 2012 | ND2 | 5108 |
| Zhuo 2012 | ND2 | 5147 |
| Zhuo 2012 | ND2 | 5178 |
| Zhuo 2012 | ND2 | 5231 |
| Zhuo 2012 | ND2 | 5263 |
| Zhuo 2012 | ND2 | 5301 |
| Zhuo 2012 | ND2 | 5417 |
| Zhuo 2012 | ND2 | 5460 |
| Ding 2018 | ND2 | 4715 |
| Ding 2018 | ND2 | 4820 |
| Zhuo 2010 | ND3 | 10310 |
| Zhuo 2010 | ND3 | 10397 |
| Zhuo 2010 | ND3 | 10398 |
| Zhuo 2010 | ND3 | 10400 |
| Zhuo 2012 | ND3 | 10097 |
| Zhuo 2012 | ND3 | 10238 |
| Zhuo 2012 | ND3 | 10310 |
| Zhuo 2012 | ND3 | 10398 |
| Zhuo 2012 | ND3 | 10400 |
| Ding 2016 | ND3 | 10398 |
| Ding 2016 | ND3 | 10400 |
| Zhuo 2012 | ND4 | 10873 |
| Zhuo 2012 | ND4 | 10915 |
| Zhuo 2012 | ND4 | 11084 |
| Zhuo 2012 | ND4 | 11335 |
| Zhuo 2012 | ND4 | 11440 |
| Zhuo 2012 | ND4 | 11719 |
| Zhuo 2012 | ND4 | 11944 |
| Zhuo 2012 | ND4 | 12026 |

| Study ID | Gene | Position |
|---|---|---|
| Zhuo 2010 | ND4 | 10609 |
| Zhuo 2010 | ND4 | 10873 |
| Zhuo 2010 | ND4 | 11084 |
| Zhuo 2010 | ND4 | 11914 |
| Zhuo 2010 | ND4 | 11944 |
| Zhuo 2010 | ND4 | 12026 |
| Ding 2016 | ND4 | 10873 |
| Ding 2016 | ND4 | 11719 |
| Ding 2018 | ND4 | 11719 |
| Zhuo 2010 | ND5 | 12406 |
| Zhuo 2010 | ND5 | 12705 |
| Zhuo 2010 | ND5 | 12882 |
| Zhuo 2010 | ND5 | 13759 |
| Zhuo 2010 | ND5 | 13824 |
| Zhuo 2010 | ND5 | 13928 |
| Zhuo 2010 | ND5 | 13942 |
| Zhuo 2010 | ND5 | 14067 |
| Zhuo 2012 | ND5 | 12406 |
| Zhuo 2012 | ND5 | 12705 |
| Zhuo 2012 | ND5 | 12811 |
| Zhuo 2012 | ND5 | 13145 |
| Zhuo 2012 | ND5 | 13263 |
| Zhuo 2012 | ND5 | 13434 |
| Zhuo 2012 | ND5 | 13708 |
| Zhuo 2012 | ND5 | 13759 |
| Zhuo 2012 | ND5 | 13928 |
| Ding 2016 | ND5 | 12705 |
| Ding 2016 | ND5 | 13928 |
| Ding 2018 | ND5 | 13928 |
| Ding 2017 | ND5 | 12811 |
| Ding 2016 | ND5 | 12338 |
| Zhuo 2010 | ND6 | 14668 |
| Zhuo 2012 | ND6 | 14199 |
| Zhuo 2012 | ND6 | 14272 |
| Zhuo 2012 | ND6 | 14365 |
| Zhuo 2012 | ND6 | 14569 |
| Zhuo 2012 | ND6 | 14668 |
| Ding 2018 | ND6 | 14470 |

#### SUPPLEMENT 3 - POLYMORPHISMS IN GENES CODING FOR MITOCHONDRIAL tRNAs

**All polymorphisms detected in genes coding for mt-tRNAs**

| Study ID | Gene | Position | PCOS sample size | PCOS with SNP | PCOS without SNP | Frequency in PCOS (%) | Control sample size | Controls with SNP | Controls without SNP | Frequency in Controls (%) |
|---|---|---|---|---|---|---|---|---|---|---|
| Zhuo 2012 | tRNA Arg | 10454 | 57 | 1 | 56 | 1.7 | 38 | 0 | 38 | 0 |
| Ding 2017 | tRNA Arg | 10454 | 80 | 2 | 78 | 2.5 | 50 | 0 | 50 | 0 |
| Zhuo 2012 | tRNA Asp | 7543 | 57 | 1 | 56 | 1.7 | 38 | 0 | 38 | 0 |
| Ding 2017 | tRNA Asp | 7543 | 80 | 1 | 79 | 1.25 | 50 | 0 | 50 | 0 |
| Zhuo 2012 | tRNA Cys | 5821 | 57 | 1 | 56 | 1.7 | 38 | 0 | 38 | 0 |
| Ding 2017 | tRNA Cys | 5821 | 80 | 3 | 77 | 3.75 | 50 | 1 | 49 | 2 |
| Zhuo 2010 | tRNA Gln | 14693 | N/A | N/A | N/A | N/A | N/A | N/A | N/A | N/A |
| Ding 2018 | tRNA Gln | 4363 | N/A | N/A | N/A | N/A | N/A | N/A | N/A | N/A |
| Ding 2017 | tRNA Gln | 4363 | 80 | 4 | 76 | 5 | 50 | 0 | 50 | 0 |
| Ding 2017 | tRNA Gln | 4395 | 80 | 1 | 79 | 1.25 | 50 | 0 | 50 | 0 |
| Zhuo 2012 | tRNA Glu | 4395 | 57 | 17 | 40 | 29.8 | 38 | 0 | 38 | 0 |
| Ding 2017 | tRNA Glu | 14693 | 80 | 1 | 79 | 1.25 | 50 | 0 | 50 | 0 |
| Ding 2017 | tRNA Met | 4454 | 80 | 0 | 80 | 0 | 50 | 2 | 48 | 4 |
| Ding 2016 | tRNA Leu | 3302 | N/A | N/A | N/A | N/A | N/A | N/A | N/A | N/A |
| Ding 2018 | tRNA Leu | 3275 | N/A | N/A | N/A | N/A | N/A | N/A | N/A | N/A |
| Ding 2017 | tRNA Leu | 3302 | 80 | 1 | 79 | 1.25 | 50 | 0 | 50 | 0 |
| Ding 2017 | tRNA Leu | 3275 | 80 | 2 | 78 | 2.5 | 50 | 0 | 50 | 0 |
| Saeed 2019 | tRNA Leu | 3157 | 70 | 2 | 68 | 2.86 | 59 | 0 | 59 | 0 |
| Saeed 2019 | tRNA Leu | 3162 | 70 | 0 | 70 | 0 | 59 | 0 | 59 | 0 |
| Saeed 2019 | tRNA Leu | 3203 | 70 | 3 | 67 | 4.29 | 59 | 1 | 58 | 1.69 |
| Saeed 2019 | tRNA Leu | 3282 | 70 | 2 | 68 | 2.86 | 59 | 0 | 59 | 0 |
| Saeed 2019 | tRNA Leu | 3285 | 70 | 2 | 68 | 2.86 | 59 | 0 | 59 | 0 |
| Saeed 2019 | tRNA Leu | 3302 | 70 | 2 | 68 | 2.86 | 59 | 0 | 59 | 0 |
| Saeed 2019 | tRNA Leu | 3275 | 70 | 4 | 66 | 5.71 | 59 | 0 | 59 | 0 |

| | | | | | | | | | | |
|---|---|---|---|---|---|---|---|---|---|---|
| Saeed 2019 | tRNA Leu | 4225 | 70 | 1 | 69 | 1.43 | 59 | 0 | 59 | 0 |
| Saeed 2019 | tRNA Leu | 5206 | 70 | 3 | 67 | 4.29 | 59 | 0 | 59 | 0 |
| Saeed 2019 | tRNA Leu | 8434 | 70 | 1 | 69 | 1.43 | 59 | 3 | 56 | 5.08 |
| Zhuo 2012 | tRNA Lys | 8343 | 57 | 1 | 56 | 1.7 | 38 | 0 | 38 | 0 |
| Ding 2018 | tRNA Lys | 8343 | N/A | N/A | N/A | N/A | N/A | N/A | N/A | N/A |
| Ding 2017 | tRNA Lys | 8343 | 80 | 2 | 78 | 2.5 | 50 | 0 | 50 | 0 |
| Ding 2017 | tRNA Ser | 7492 | 80 | 1 | 79 | 1.25 | 50 | 0 | 50 | 0 |
| Ding 2016 | tRNA Ser | 7492 | N/A | N/A | N/A | N/A | N/A | N/A | N/A | N/A |
| Ding 2017 | tRNA Thr | 15900 | 80 | 0 | 80 | 0 | 50 | 1 | 49 | 2 |

**Dataset used in meta-analysis**

| Mitochondrial-encoded tRNA | Frequency of SNP in PCOS (%) | Frequency of SNP in Control (%) |
|---|---|---|
| Cys | 2.9 | 1.1 |
| Leu | 2.7 | 0.6 |
| Glu | 13.1 | 0.0 |
| Gln | 3.1 | 0.0 |
| Lys | 2.2 | 0.0 |
| Arg | 1.9 | 0.0 |
| Asp | 1.5 | 0.0 |

#### SUPPLEMENT 4 – POLYMORPHISMS IN GENES CODING FOR MITOCHONDRIAL rRNAs

| Study ID | Gene | Position |
|---|---|---|
| Zhuo 2010 | 12S rRNA | 663 |
| Zhuo 2010 | 12S rRNA | 709 |
| Zhuo 2010 | 12S rRNA | 752 |
| Zhuo 2010 | 12S rRNA | 827 |
| Zhuo 2010 | 12S rRNA | 1438 |
| Zhuo 2012 | 12S rRNA | 663 |
| Zhuo 2012 | 12S rRNA | 709 |
| Zhuo 2012 | 12S rRNA | 750 |
| Zhuo 2012 | 12S rRNA | 752 |
| Zhuo 2012 | 12S rRNA | 827 |
| Zhuo 2012 | 12S rRNA | 1382 |
| Zhuo 2012 | 12S rRNA | 1438 |
| Ding 2016 | 12S rRNA | 750 |
| Ding 2016 | 12S rRNA | 1438 |
| Ding 2018 | 12S rRNA | 750 |
| Ding 2018 | 12S rRNA | 827 |
| Ding 2018 | 12S rRNA | 1438 |
| Zhuo 2010 | 16S rRNA | 1736 |
| Zhuo 2010 | 16S rRNA | 3010 |
| Zhuo 2012 | 16S rRNA | 1736 |
| Zhuo 2012 | 16S rRNA | 2706 |
| Zhuo 2012 | 16S rRNA | 3010 |
| Ding 2016 | 16S rRNA | 1734 |
| Ding 2016 | 16S rRNA | 2706 |
| Ding 2018 | 16S rRNA | 2706 |
| Ding 2018 | 16S rRNA | 3109 |

#### SUPPLEMENT 5 – POLYMORPHISMS IN THE D-LOOP OF THE MITOCHONDRIAL GENOME

##### All polymorphisms detected in the D-Loop of the mitochondrial genome

| Study ID | Polymorphism | PCOS sample size | PCOS with SNP | PCOS without SNP | Frequency in PCOS (%) | Control sample size | Control with SNP | Control without SNP | Frequency in controls (%) |
|---|---|---|---|---|---|---|---|---|---|
| Zhuo 2012 | A73G | 57 | 10 | 47 | 17.5 | 38 | 8 | 30 | 21.6 |
| Zhuo 2012 | T146C | 57 | 1 | 56 | 1.7 | 38 | 1 | 37 | 2.7 |
| Zhuo 2012 | C150T | 57 | 6 | 51 | 10.5 | 38 | 3 | 35 | 8.1 |
| Zhuo 2012 | A153G | 57 | 4 | 53 | 7.0 | 38 | 2 | 36 | 5.4 |
| Zhuo 2012 | T195A | 57 | 5 | 52 | 8.7 | 38 | 2 | 36 | 5.4 |
| Zhuo 2012 | A235G | 57 | 3 | 54 | 5.3 | 38 | 2 | 36 | 5.4 |
| Zhuo 2012 | A263G | 57 | 8 | 49 | 14.0 | 38 | 7 | 31 | 18.9 |
| Zhuo 2012 | G316A | 57 | 1 | 56 | 1.7 | 38 | 0 | 38 | 0 |
| Zhuo 2012 | T489A | 57 | 4 | 53 | 7.0 | 38 | 3 | 35 | 8.1 |
| Zhuo 2012 | G16129A | 57 | 2 | 55 | 3.5 | 38 | 2 | 36 | 5.4 |
| Zhuo 2012 | T16140C | 57 | 3 | 54 | 5.3 | 38 | 0 | 38 | 0 |
| Zhuo 2012 | T16189C | 57 | 6 | 51 | 10.5 | 38 | 4 | 34 | 10.8 |
| Zhuo 2012 | C16223T | 57 | 9 | 48 | 15.8 | 38 | 2 | 36 | 5.4 |
| Zhuo 2012 | A16316G | 57 | 6 | 51 | 10.5 | 38 | 5 | 33 | 13.5 |
| Zhuo 2012 | T16357C | 57 | 1 | 56 | 1.7 | 38 | 0 | 38 | 0 |
| Zhuo 2012 | T16519C | 57 | 2 | 55 | 3.5 | 38 | 1 | 37 | 2.7 |
| Reddy 2019 | A73G | 118 | 75 | 43 | 63.6 | 114 | 75 | 39 | 65.8 |
| Reddy 2019 | A93G | 118 | 7 | 111 | 5.9 | 114 | 4 | 110 | 3.5 |
| Reddy 2019 | T146C | 118 | 8 | 110 | 6.8 | 114 | 12 | 102 | 10.5 |
| Reddy 2019 | T152C | 118 | 23 | 95 | 19.5 | 114 | 23 | 91 | 20.2 |
| Reddy 2019 | A189G | 118 | 31 | 87 | 26.3 | 114 | 15 | 99 | 13.2 |
| Reddy 2019 | T195A | 118 | 8 | 110 | 6.8 | 114 | 7 | 107 | 6.1 |
| Reddy 2019 | A263G | 118 | 72 | 46 | 61.0 | 114 | 76 | 38 | 66.7 |
| Reddy 2019 | 316C | 118 | 71 | 47 | 60.1 | 114 | 68 | 46 | 59.6 |
| Reddy 2019 | T489C | 118 | 25 | 93 | 21.2 | 114 | 14 | 100 | 12.3 |
| Reddy 2019 | 522CA | 118 | 15 | 103 | 12.7 | 114 | 7 | 107 | 6.1 |

| | | | | | | | | | |
|---|---|---|---|---|---|---|---|---|---|
| Reddy 2019 | G16129A | 118 | 9 | 109 | 7.6 | 114 | 11 | 103 | 9.6 |
| Reddy 2019 | T16172C | 118 | 10 | 108 | 8.5 | 114 | 11 | 103 | 9.6 |
| Reddy 2019 | T16189C | 118 | 14 | 104 | 11.9 | 114 | 15 | 99 | 13.2 |
| Reddy 2019 | C16223T | 118 | 44 | 74 | 37.3 | 114 | 52 | 62 | 45.6 |
| Reddy 2019 | T16311C | 118 | 13 | 105 | 11.0 | 114 | 16 | 98 | 14.0 |
| Reddy 2019 | G16319A | 118 | 12 | 106 | 10.2 | 114 | 5 | 109 | 4.4 |
| Reddy 2019 | T16362C | 118 | 14 | 104 | 11.9 | 114 | 8 | 106 | 7.0 |
| Reddy 2019 | T16519C | 118 | 63 | 55 | 53.4 | 114 | 54 | 60 | 47.4 |
| Reddy 2019 | D310 | 118 | 64 | 54 | 54.2 | 114 | 33 | 81 | 28.9 |
| Deng 2021 | C150T | 421 | 79 | 342 | 18.76 | 409 | 103 | 306 | 25.18 |
| Deng 2021 | G207A | 421 | 11 | 410 | 2.61 | 409 | 28 | 381 | 6.85 |
| Deng 2021 | A263G | 421 | 418 | 3 | 99.29 | 409 | 399 | 10 | 97.56 |
| Deng 2021 | 16036G | 421 | 24 | 397 | 5.70 | 409 | 42 | 367 | 10.27 |
| Deng 2021 | 16036GG | 421 | 2 | 419 | 0.48 | 409 | 22 | 387 | 5.38 |
| Deng 2021 | 16049G | 421 | 2 | 419 | 0.48 | 409 | 36 | 373 | 8.80 |
| Deng 2021 | C16234T | 421 | 17 | 404 | 4.04 | 409 | 30 | 379 | 7.33 |
| Deng 2021 | T16362C | 421 | 169 | 252 | 40.14 | 409 | 196 | 213 | 47.92 |
| Hu 2011 | 16094T/C | 77 | 7 | 70 | 9.09 | 45 | 4 | 41 | 8.89 |
| Hu 2011 | C16173T | 77 | 67 | 10 | 87.01 | 45 | 38 | 7 | 84.40 |
| Hu 2011 | 16181 – 16195 | 77 | 76 | 1 | 98.70 | 45 | 44 | 1 | 97.78 |
| Hu 2011 | A2C12 | 77 | 6 | 71 | 7.89 | 45 | 1 | 44 | 2.27 |
| Hu 2011 | A3C11 | 77 | 9 | 68 | 11.84 | 45 | 8 | 37 | 18.18 |
| Hu 2011 | A4C9T | 77 | 48 | 29 | 63.16 | 45 | 31 | 14 | 70.45 |
| Hu 2011 | A4C8T2 | 77 | 13 | 64 | 17.11 | 45 | 4 | 41 | 9.09 |
| Hu 2011 | T16225C | 77 | 24 | 53 | 31.17 | 45 | 14 | 31 | 31.11 |
| Hu 2011 | T16300C | 77 | 9 | 68 | 11.69 | 45 | 8 | 37 | 17.78 |
| Hu 2011 | T16306C | 77 | 10 | 67 | 12.99 | 45 | 4 | 41 | 8.89 |
| Hu 2011 | T16313C | 77 | 10 | 67 | 12.99 | 45 | 7 | 38 | 15.56 |
| Hu 2011 | G16321A | 77 | 9 | 68 | 11.69 | 45 | 8 | 37 | 17.78 |
| Hu 2011 | T16364C | 77 | 28 | 49 | 36.36 | 45 | 18 | 27 | 40.00 |

| | | | | | | | | | |
|---|---|---|---|---|---|---|---|---|---|
| Hu 2011 | T146C | 77 | 9 | 68 | 11.69 | 45 | 5 | 40 | 11.11 |
| Hu 2011 | C150T | 77 | 62 | 15 | 80.52 | 45 | 39 | 6 | 80.00 |
| Hu 2011 | T152C | 77 | 19 | 58 | 24.68 | 45 | 9 | 36 | 20.00 |
| Hu 2011 | C195T | 77 | 66 | 11 | 85.71 | 45 | 42 | 3 | 93.33 |
| Hu 2011 | A248 | 77 | 17 | 60 | 22.08 | 45 | 9 | 36 | 20.00 |
| Hu 2011 | T491C | 77 | 48 | 29 | 62.34 | 45 | 22 | 23 | 48.89 |
| Hu 2011 | 303-317 | 77 | 76 | 1 | 98.70 | 45 | 44 | 1 | 97.78 |
| Hu 2011 | C8TC6 | 77 | 41 | 36 | 53.95 | 45 | 18 | 27 | 40.91 |
| Hu 2011 | C9TC6 | 77 | 11 | 66 | 14.47 | 45 | 5 | 40 | 11.36 |
| Hu 2011 | C7TC6 | 77 | 24 | 53 | 31.58 | 45 | 21 | 24 | 47.73 |
| Zhuo 2010 | T146C | - | - | - | - | - | - | - | - |
| Zhuo 2010 | T150C | - | - | - | - | - | - | - | - |
| Zhuo 2010 | T152C | - | - | - | - | - | - | - | - |
| Zhuo 2010 | C195T | - | - | - | - | - | - | - | - |
| Zhuo 2010 | A200G | - | - | - | - | - | - | - | - |
| Zhuo 2010 | G207A | - | - | - | - | - | - | - | - |
| Zhuo 2010 | A235G | - | - | - | - | - | - | - | - |
| Zhuo 2010 | A263G | - | - | - | - | - | - | - | - |
| Zhuo 2010 | T489C | - | - | - | - | - | - | - | - |
| Zhuo 2010 | G16129A | - | - | - | - | - | - | - | - |
| Zhuo 2010 | T16172C | - | - | - | - | - | - | - | - |
| Zhuo 2010 | T16189C | - | - | - | - | - | - | - | - |
| Zhuo 2010 | C16223T | - | - | - | - | - | - | - | - |
| Zhuo 2010 | C16290T | - | - | - | - | - | - | - | - |
| Zhuo 2010 | G16319A | - | - | - | - | - | - | - | - |
| Zhuo 2010 | T16362C | - | - | - | - | - | - | - | - |
| Zhuo 2010 | T16519C | - | - | - | - | - | - | - | - |
| Ding 2016 | A73G | - | - | - | - | - | - | - | - |
| Ding 2016 | T152C | - | - | - | - | - | - | - | - |
| Ding 2016 | D310 | - | - | - | - | - | - | - | - |

| | | | | | | | | | |
|---|---|---|---|---|---|---|---|---|---|
| Ding 2016 | T16189C | - | - | - | - | - | - | - | - |
| Ding 2016 | T16519C | - | - | - | - | - | - | - | - |
| Ding 2018 | A73G | - | - | - | - | - | - | - | - |
| Ding 2018 | C150T | - | - | - | - | - | - | - | - |
| Ding 2018 | C328T | - | - | - | - | - | - | - | - |
| Ding 2018 | T16142C | - | - | - | - | - | - | - | - |
| Ding 2018 | T16189C | - | - | - | - | - | - | - | - |
| Ding 2018 | T16224C | - | - | - | - | - | - | - | - |

#### Dataset used in meta-analysis

| **Polymorphism** | **Study ID** | **PCOS sample size** | **PCOS with SNP** | **PCOS without SNP** | **Frequency in PCOS (%)** | **Control sample size** | **Control with SNP** | **Control without SNP** | **Frequency in controls (%)** |
|---|---|---|---|---|---|---|---|---|---|
| C150T | Hu 2011 | 77 | 62 | 15 | 80.52 | 45 | 39 | 6 | 80.00 |
| | Zhuo 2012 | 57 | 6 | 51 | 10.5 | 38 | 3 | 35 | 8.1 |
| | Deng 2021 | 421 | 79 | 342 | 18.76 | 409 | 103 | 306 | 25.18 |
| T146C | Hu 2011 | 77 | 9 | 68 | 11.69 | 45 | 5 | 40 | 11.11 |
| | Zhuo 2012 | 57 | 1 | 56 | 1.7 | 38 | 1 | 37 | 2.7 |
| | Reddy 2019 | 118 | 8 | 110 | 6.8 | 114 | 12 | 102 | 10.5 |
| A263G | Zhuo 2012 | 57 | 8 | 49 | 14.0 | 38 | 7 | 31 | 18.9 |
| | Reddy 2019 | 118 | 72 | 46 | 61.0 | 114 | 76 | 38 | 66.7 |
| | Deng 2021 | 421 | 418 | 3 | 99.29 | 409 | 399 | 10 | 97.56 |

#### SUPPLEMENT 6 – MtDNA COPY NUMBERS

| | Study ID | Cells | Total Sample Size (n) | PCOS Sample Size | Control Sample Size | PCOS Log (mean mt CN) | Control Log (mean mt CN) | PCOS SEM | Control SEM | PCOS SD | Control SD |
|---|---|---|---|---|---|---|---|---|---|---|---|
| Included in meta-analysis | Ding et al. 2017 | Blood | 130 | 80 | 50 | 0.29 | 0.84 | 0.31 | 0.88 | 0.44 | 0.79 |
| | Reddy et al. 2019 | Blood | 232 | 118 | 114 | 1.36 | 1.50 | 0.44 | 0.56 | 4.78 | 5.98 |
| | Shukla et al. 2020 | Blood | 60 | 30 | 30 | 1.22 | 1.64 | 0.33 | 0.35 | 1.81 | 1.92 |
| | Wang et al. 2020 | Granulosa | 107 | 39 | 68 | 0.761 | 1.021 | 0.05 | 0.06 | 0.31 | 0.49 |
| Insufficient data for meta-analysis | Lee et al. 2011 | Blood | 110 | 50 | 60 | 1.44 | 1.88 | - | - | - | - |
| | Ding et al. 2016 | Blood | - | - | - | - | - | - | - | - | - |
| | Ding et al. 2018 | Blood | - | - | - | - | - | - | - | - | - |
| | Saeed et al. 2019 | Blood | 63 | 38 | 25 | 0.88 | 1.00 | - | - | - | - |
| | Yang et al. 2020 | Blood | 88 | 0 | - | - | - | - | - | - | - |

#### SUPPLEMENT 7 – QUALITY ASSESSMENT SCORES

| Study ID | Rationale for study | Selection and definition of outcome of interest | Selection and comparability of comparison group | Technical classification of the exposure | Non-technical classification of the exposure | Other sources of bias | Sample size and power | A priori planning of analyses | Statistical methods and control for confounding | Testing of assumptions and inferences for genetic analyses | Appropriateness of inferences drawn from results | Total score | Quality status |
|---|---|---|---|---|---|---|---|---|---|---|---|---|---|
| Zhuo 2010 | 5 | 4 | 4 | 3 | 1 | 1 | 2 | 2 | 1 | 3 | 3 | 29 | Poor |
| Hu 2011 | 4 | 3 | 4 | 5 | 3 | 3 | 2 | 5 | 6 | 4 | 3 | 42 | Moderate |
| Lee 2011 | 6 | 5 | 6 | 6 | 5 | 7 | 2 | 5 | 6 | 6 | 6 | 60 | Good |
| Rabol 2011 | 7 | 4 | 5 | 3 | 2 | 1 | 1 | 5 | 3 | 4 | 5 | 40 | Moderate |
| Zhuo 2012 | 6 | 5 | 5 | 3 | 3 | 2 | 2 | 3 | 5 | 5 | 4 | 43 | Moderate |
| Ding 2016a | 2 | 2 | 2 | 2 | 1 | 2 | 1 | 2 | 1 | 3 | 1 | 19 | Poor |
| Ding 2016b | 1 | 2 | 1 | 1 | 1 | 1 | 1 | 2 | 1 | 1 | 1 | 13 | Poor |
| Ding 2017 | 6 | 6 | 6 | 6 | 6 | 2 | 3 | 6 | 4 | 7 | 6 | 58 | Good |
| Ding 2018 | 3 | 2 | 2 | 3 | 1 | 2 | 1 | 3 | 2 | 4 | 2 | 25 | Poor |
| Reddy 2019 | 7 | 6 | 7 | 7 | 7 | 3 | 3 | 7 | 7 | 6 | 3 | 63 | Good |
| Saeed 2019 | 4 | 2 | 4 | 6 | 4 | 3 | 2 | 6 | 5 | 1 | 2 | 39 | Moderate |
| Yang 2020 | 7 | 2 | - | 6 | 3 | 6 | 2 | 7 | 5 | 5 | 6 | 49 | Good |
| Shukla 2020 | 6 | 3 | 6 | 6 | 3 | 1 | 2 | 5 | 2 | 5 | 5 | 44 | Moderate |
| Wang 2020 | 5 | 4 | 4 | 7 | 3 | 1 | 1 | 4 | 3 | 5 | 5 | 42 | Moderate |
| Deng 2021 | 7 | 3 | 5 | 3 | 3 | 1 | 4 | 7 | 7 | 6 | 4 | 50 | Good |
| Median | 6 | 3 | 4 | 5 | 3 | 2 | 2 | 5 | 4 | 5 | 4 | | |
| Q1 | 4 | 2 | 2 | 3 | 1 | 1 | 1 | 3 | 2 | 3 | 2 | | |
| Q3 | 7 | 5 | 6 | 6 | 4 | 3 | 2 | 6 | 6 | 6 | 5 | | |
| IQR | 4-7 | 2-5 | 2-6 | 3-6 | 1-4 | 1-3 | 1-2 | 3-6 | 2-6 | 3-6 | 2-5 | | |
